# The Prevalence of ocular manifestations and ocular samples polymerase chain reaction positivity in patients with COVID 19 - a systematic review and meta-analysis

**DOI:** 10.1101/2020.06.29.20142414

**Authors:** Soumen Sadhu, Sushmitha Arcot Dandapani, Deepmala Mazumdar, Sangeetha Srinivasan, Jyotirmay Biswas

## Abstract

**Objective:** To estimate the prevalence of ocular manifestations and ocular samples polymerase chain reaction (PCR) positivity among COVID-19 patients.

**Methods:** A systematic literature review was performed using search engines (PubMed, Google Scholar, Medrixv and BioRixv) with keywords “SARS-CoV-2”, “novel coronavirus”, “COVID-19”, “ocular manifestations”, “conjunctival congestion”, “Ocular detection”, “Polymerase chain reaction”, and “conjunctivitis”. The measure of heterogeneity was evaluated with the I^2^ statistic. The pooled proportion of patients presenting with symptoms and ocular samples PCR positivity was estimated.

**Results:** A total of 20 studies (14 studies and 6 case-reports) were included in the systematic review and 14 studies were included in the meta-analysis. The pooled prevalence of conjunctivitis was 5.17% (95% CI: 2.90-8.04). Conjunctivitis was reported as an initial symptom of the disease in 0.858 % (95% CI: 0.31-1.67). Common associated features include itching, chemosis, epiphora. Seven patients (29 %) with conjunctivitis showed positive results in ocular samples, whereas 13 patients (54%) showed positive only in their *nasopharyngeal samples (NPs) or* sputum samples and 4 patients (16 %) were negative for both NPs and Sputum as well as ocular samples. The pooled prevalence of ocular PCR positivity was 2.90 % (95% CI: 1.77 – 4.46) vs. NPs 89.8% (95% CI: 78.80-79.0).

**Conclusion:** The prevalence of conjunctivitis and ocular samples PCR positivity among COVID-19 patients was low indicating that the eye is a less affected organ. However, conjunctivitis may present as the first symptom of the disease making the patient seek medical care at the earliest.

**Synopsis:** Viral conjunctivitis was the only symptom reported. The prevalence of conjunctivitis and ocular samples polymerase chain reaction positivity among COVID-19 patients was low indicating the eye is a less effected organ.

## Introduction

Coronaviruses are single-stranded positive-sense ribonucleic acid (RNA) viruses known to cause upper respiratory tract infections. Apart from the previously known strains of human corona-viruses (HCoV −229E, HCoV-NL63, HCoV-OC43, HCoV-HKU1, SARS-CoV, MERS-CoV),^1^ a new strain was identified to cause severe acute respiratory syndrome (SARS) in January 2020 and named as SARS-CoV-2 and the disease as the novel coronavirus disease-2019 (COVID-19).^2^

The novel SARS-CoV-2 has a close genetic sequential identity^3^ to SARS CoV-1, hence the SARS-CoV-1 pandemic is used as a model to predict the behavior of SARS-CoV-2. After the outbreak, COVID-19 gained pandemic proportions within a very short time killing millions of people worldwide. The SARS-CoV-2 virus is believed to be transmitted through inhalation of infected respiratory droplets or touching contaminated surfaces, which can in turn spread through the mucous membranes of the mouth, eyes, and nose (routes of various microbial transmissions).^4^The binding cellular receptor, angiotensin-converting enzyme 2 (ACE2) for SARS-CoV is extensively expressed in the respiratory tract, lung alveoli, and intestinal epithelial cells.ACE2 receptors were found on the corneal, conjunctival epithelium, aqueous and vitreous humor, although to a lesser extent as compared to respiratory tissues.^5,6^Due to the low expressions of ACE2 receptors in the ocular surface, the viral load is reported to be low or undetectable in the polymerase chain reaction (PCR) assay.^5^

In experimental animal models of rhesus macaques, when inoculated with infectious doses of SARS-CoV-2 though the conjunctival sac, they became potentially infected with the virus and developed interstitial pneumonia.^7^ Similarly, sight-threatening conditions like granulomatous anterior uveitis, retinal vasculitis, choroiditis, retinal detachment, virus-induced macular degeneration, and optic neuritis were also reported in cats infected with the feline (feline infectious peritonitis) and murine coronavirus (mouse hepatitis virus).^1^ The above findings suggest the possibility of the conjunctiva and exposed ocular surfaces a gateway forSARS-CoV-2 infection and transmission leading to a respiratory involvement. In this context, there is an increased interest in evaluating ocular manifestations and the presence of the virus in ocular secretions in patients infected with the novel coronavirus.

Ocular manifestations and the presence of the viral RNA in ocular secretions have been reported by several observational and interventional studies. It has been postulated that conjunctivitis can be the first presenting symptom of the disease.^1^ Nevertheless, the ocular presentations or manifestations of the virus in affected patients are not well understood.

The goal of the study is to perform a systematic review and meta-analysis to provide a comprehensive outlook of the prevalence of Ocular manifestations in patients suffering from COVID-19. The main objectives of the study were i) To estimate the prevalence of ocular signs and symptoms among COVID-19 patients, its onset, duration and prognosis ii) To estimate the proportion of patients presenting with conjunctivitis as a first symptom of the disease iii) To estimate the proportion of patients having ocular sample PCR positivity.

## Materials and methods

### Inclusion criteria

We included all published/pre-prints papers and case reports which aimed at evaluating ocular manifestations and PCR results of ocular samples in patients with COVID-19. We included the reported observational studies for meta-analysis, the inclusion criteria were i) studies aimed at looking into ocular manifestations among COVID-19 patients, either clinically or laboratory-confirmed (Nasopharyngeal samples (NPs) / Sputum PCR or radiographic investigations) cases. ii) Studies that evaluated ocular samples by PCR assay for the detection of viral RNA. We reserved all case reports only for the systematic review purpose. The literature search included only those articles published in English and those for which English translation was readily available on the journal site (through Google translate). There were no restrictions on the study population or the region.

### Literature search strategy, screening and data extraction

A comprehensive literature search was performed using four search engines PubMed, Google scholar, BioRixv and Medrixv till 10^th^ June 2020.The keywords used for the literature search were “SARS-CoV-2”, “novel coronavirus”, “COVID-19”, “ocular manifestations”, “conjunctival congestion”, “Ocular detection”, “Polymerase chain reaction”, and “conjunctivitis”,using bullion characters “AND”, “OR”, “NOT”, and the vocabulary search strategy (MeSH terms). A full-text assessment was carried out for eligible studies as per our inclusion criteria after reviewing the title and abstracts. We used the snowballing method to the references of the retrieved papers to find out any relevant articles.

Two independent investigators (SS)and (SA) reviewed the articles, performed data extraction including author, type of study, sample size, age, and the number of patients with NPs/sputum positive, duration of illness, ocular manifestations and duration, PCR results of ocular sample with and without conjunctivitis, time of ocular sample collection, ocular management and the recovery time. The extracted data were recorded in a data extraction template provided by the Cochrane library, which was then customized for our study.

The quality assessment tool for case-series studies developed by National Institutes of Health (NIH) was used for quality assessment. Each study was scored between 0 to 9. The risk of bias was classified as low (≥ 7), moderate (5-6) and high (≤4).

The data were entered in the Microsoft Excel sheet; any disagreements were resolved by further discussions among the investigators. After verification, the data were entered into the Med Calc Software (MedCalc Statistical Software version 19.3.1 (MedCalc Software Ltd, Ostend, Belgium; https://www.medcalc.org; 2020).Results were reported according to the Preferred Reporting Items for Systematic Reviews and Meta-Analyses (PRISMA) statement.^8^

### Statistical analysis

MedCalc statistical software was utilized for meta-analysis. The mean ± standard deviation was used to represent continuous data. The mean difference and 95% confidence intervals were estimated. We analyze the prevalence of conjunctivitis and PCR positivity in NPs as well as ocular. Heterogeneity was evaluated with the I^2^ statistic, in which an I^2^ value over 50% was considered an indicator of substantial heterogeneity. In the presence of heterogeneity, the random-effects model was chosen; otherwise, the fixed-effects model was applied. A funnel plot was used to investigate the publication bias. The value of 0.05 or less was considered statistically significant.

## Results

Our database search produced 1152 studies from PubMed, Google scholar, BioRixv and Medrixv. After excluding the duplicates, the abstracts for 643 articles were screened. Based on our study eligibility criteria, we included 14 studies for the meta-analysis, of which 2 were retrospective cross-sectional studies and 12 were prospective cross-sectional studies. The sample sizes of these studies ranged from 17-534 patients. We included 6 case reports for the systematic review purpose. Figure 1 shows the PRISMA flow diagram of the included studies.

**Figure 1.**
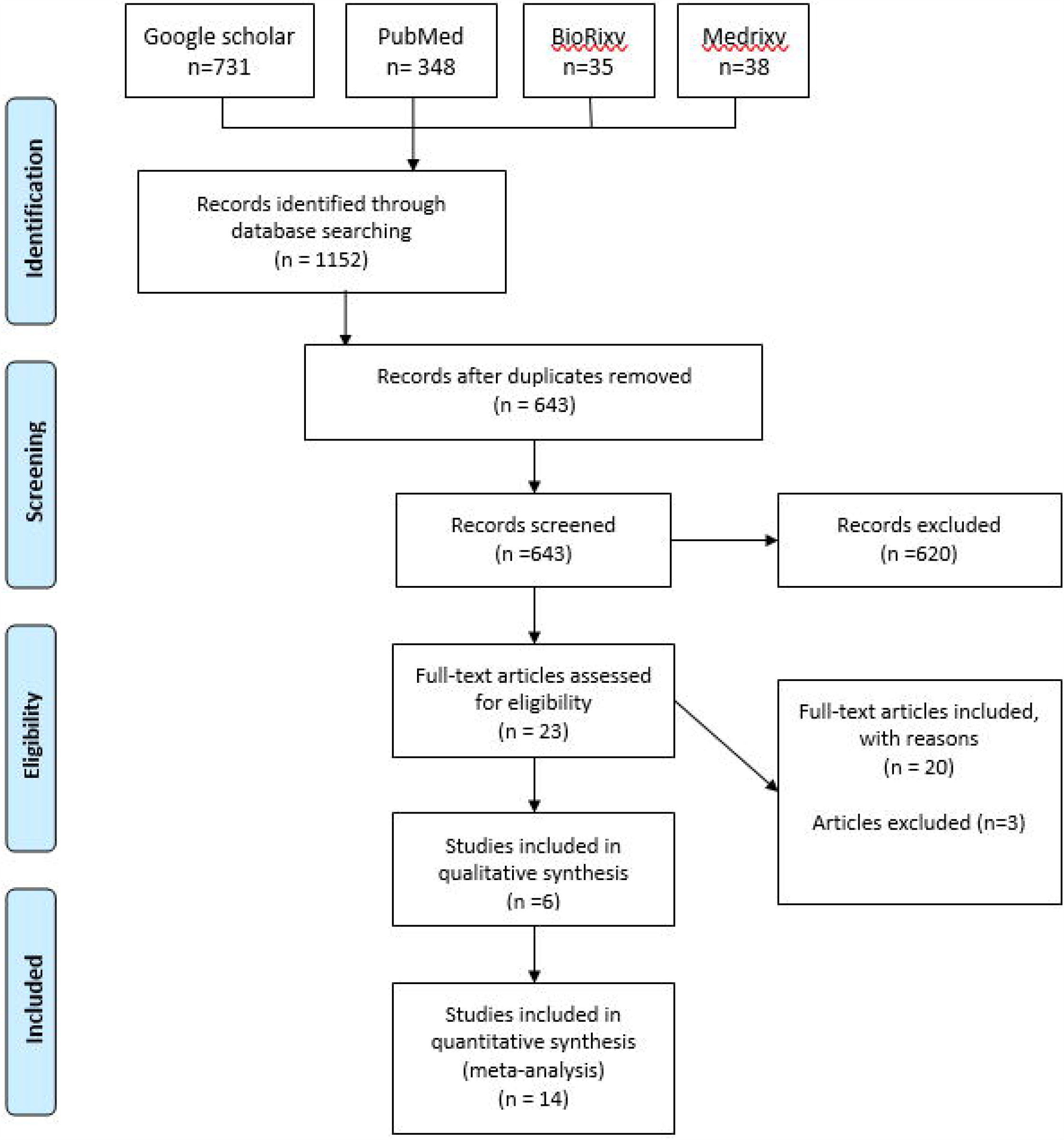
represents the PRISMA flow diagram of the included studies.

### Ocular signs and symptoms

The prevalence of ocular signs and symptoms in patients with SARS-CoV-2 was reported by 11 (sample size: 1045 patients) out of 14 studies (sample size: 1257 patients), of which, 10 studies reported the incidence of conjunctivitis. There was significant heterogeneity (I^2^= 60.27 %, p=0.007) between studies, thus we used the random effect model. The pooled prevalence estimates of conjunctivitis or conjunctival congestion were 5.17% (49 patients, 95% CI: 2.90-8.04). The forest plot with the estimated prevalence for each study has been presented in Figure 2A. The most common associated features along with conjunctivitis include epiphoria, foreign body sensation, chemosis, and itching. A detailed outline of ocular symptoms has been presented in table 1. No significant publication bias was observed between studies in the funnel plot as the maximum number of studies lies on the vertical line (Figure 2A).

**Table 1:** Characteristics of the included studies in the meta-analysis

| Author | Study Type | Sample size | Conjunctivitis (n) | Other Ocular symptoms | Total n (%) | PCR Results |  |
| --- | --- | --- | --- | --- | --- | --- | --- |
|  |  |  |  |  |  | NPs/Sputum | Ocular (%) |
| *Xia et al<sup>9</sup>* | Prospective interventional case series | 30 Confirmed NCP patients | 1/30 | **Associated Symptoms with conjunctivitis include:** Itching | 1 (3.3) | 30 | 1 (3.3) |
| *Wu et al<sup>10</sup>* | Observational case-series | 38 Clinically confirmed COVID-19 patients | 10/38 | **Associated Symptoms with conjunctivitis include:** epiphora (7, 70%), secretion (6, 60%) and Chemosis (8, 80%)<br>**Symptoms without conjunctivitis:** Epiphora 1 patient and 1 patient secretion | 12 (31.6)<br>Aggregate symptoms= epiphora 8 (21), Secretion 7 (18.4) and Chemosis 8 (21) | 28 | 2 (5.2) |
| *Yu Jun et al<sup>11</sup>* | Prospective observational | 17 COVID-19 patients | 1/17 | **Associated Symptoms with conjunctivitis include:** Chemosis | 1 (5.8) | 17 | None |
| *Xu et al<sup>12</sup>* | Retrospective cross-sectional | 30 (15 confirmed and 15 suspected cases) | NR | 1 patient only with eye itching | 1 (3.3) | 15 | None |
| *Chen et al<sup>13</sup>* | Cross-sectional prospective | 534 COVID-19 cases | 25/534 | **Associated Symptoms with conjunctivitis include:** itching (4, 16%), tearing (5, 20%), secretion (7, 28%) , FB sensation (4, 16% ) , dry eye (9, 36%).<br>**Symptoms without conjunctivitis:** Itching (49, 9.6%), tearing (50, 9.8%), secretion (45, 8.8% ), FB sensation (59, 11.6% ), dry eye (103, 20% ) | Conjunctivitis = 25 (4.7)<br>Aggregate symptoms= Itching 53 (9.9), Tearing 55 (10.3), secretion 52, (9.7), FB sensation 63 (11.8) , dry eye 103 (19.8 ) | 371 | Not done |
| *Xie et al*<sup>14</sup> | Retrospective case-series | 33 COVID-19 patients | NR | - | - | 33 | 2 (6) |
| *Sun et al*<sup>15</sup> | Cross-sectional | 102 COVID-19 patients | 2/102 | **Associated Symptoms with conjunctivitis include:** tearing | 2 (1.96) | 72 | 1 (1) |
| *Zhou et al*<sup>16</sup> | Retrospective cohort | 67 (63 laboratory confirmed and 4 suspected cases) | 1/67 | **Associated Symptoms with conjunctivitis include:** tearing and itching | 1 (1.5) | 63 | 3 (4.5) |
| *Deng et al*<sup>17</sup> | Cross-sectional observational | 114 COVID-19 pneumonia | NR | - | - | 90 | None |
| *Yan et al*<sup>18</sup> | Cross-sectional case series | 35 Confirmed cases | NR | - | - | 19 | 3 (8.5) |
| *Hong et al*<sup>19</sup> | Observational Questionnaire-based | 56 recovered discharged COVID-19 patients | 2/56 | **Associated Symptoms with conjunctivitis include:** pain and FB sensation<br>**Symptoms without conjunctivitis:** Secretion (2) , dry eyes (5), itching (3), floaters(1), FB sensation (2) | 15 (27) | 56 | 1 (1.8) |
| *Zhou et al*<sup>20</sup> | Cross-sectional observational | 121 confirmed COVID-19 patients | 3/121 | Itching (5, 65.5), redness (3, 37.5%), tearing (3, 37.5%), discharge (2, 25%), and foreign Body sensation (2, 25%). | 8 (6.6)<br>3 with conjunctivitis and 5 with other symptoms, | 121 | 3 (2.5) |
| *Liang & Wu*<sup>21</sup> | observational | 37 COVID-19 cases | 3/37 | NR | 3 (8 ) | 37 | 1 (2.7) |
| *Karimi et al*<sup>22</sup> | Prospective interventional case series | 43 Severe COVID-19 cases | 1/43 | **Associated Symptoms with conjunctivitis include: 1**<br>**Symptoms without conjunctivitis:**Foreign body sensation in 1 patient | 2 (4.65) | 30 | 3 (7) |

FB: Foreign body; NR: Not reported; NCP: New Coronavirus Pneumonia; PCR: Polymerase chain reaction; NPs: Nasopharyngeal samples

**Figure 2A.**
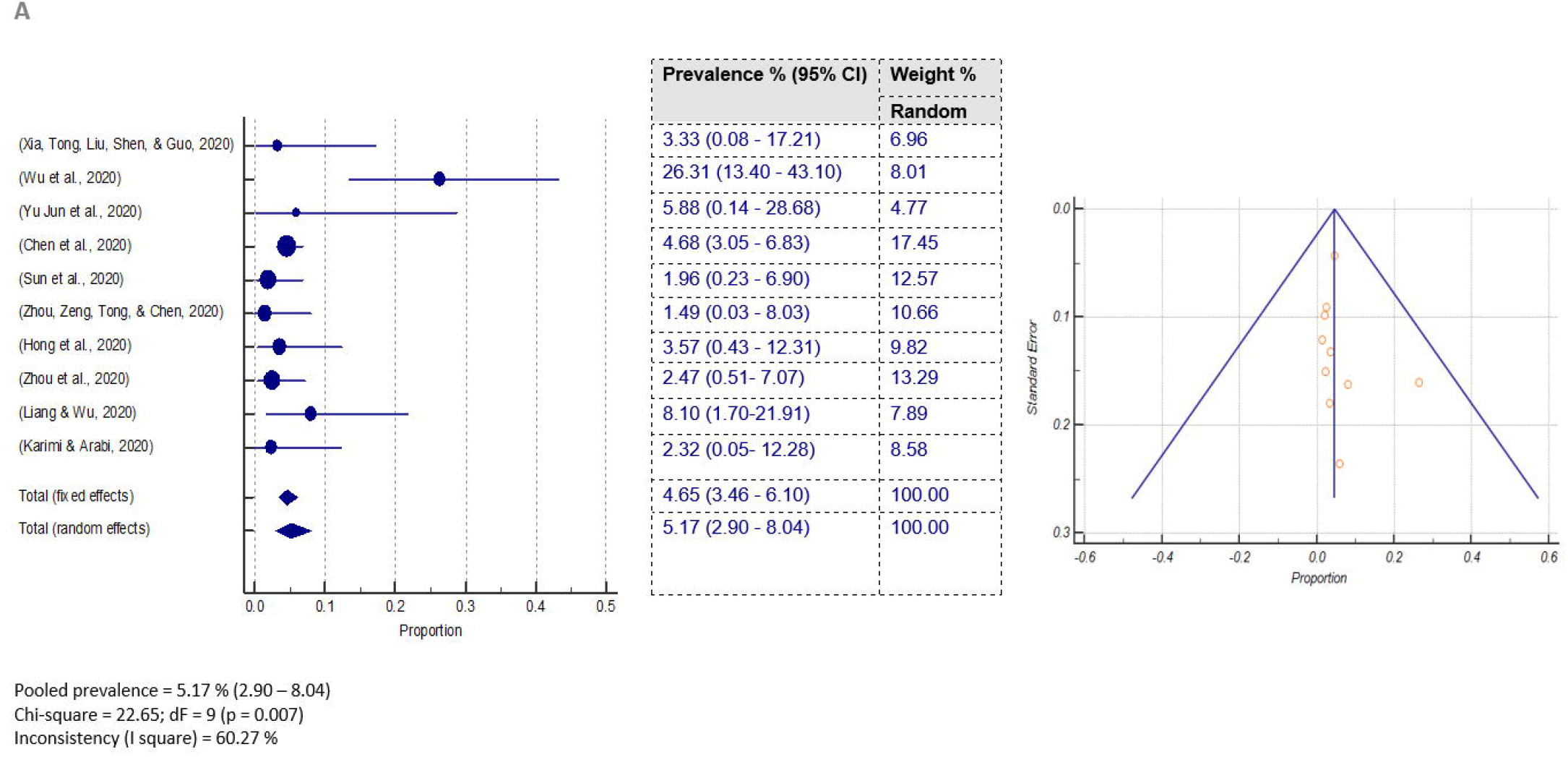
represents the forest plot (left) and the Funnel plot (right) of estimated pooled prevalence of conjunctivitis.

**Figure 2B.**
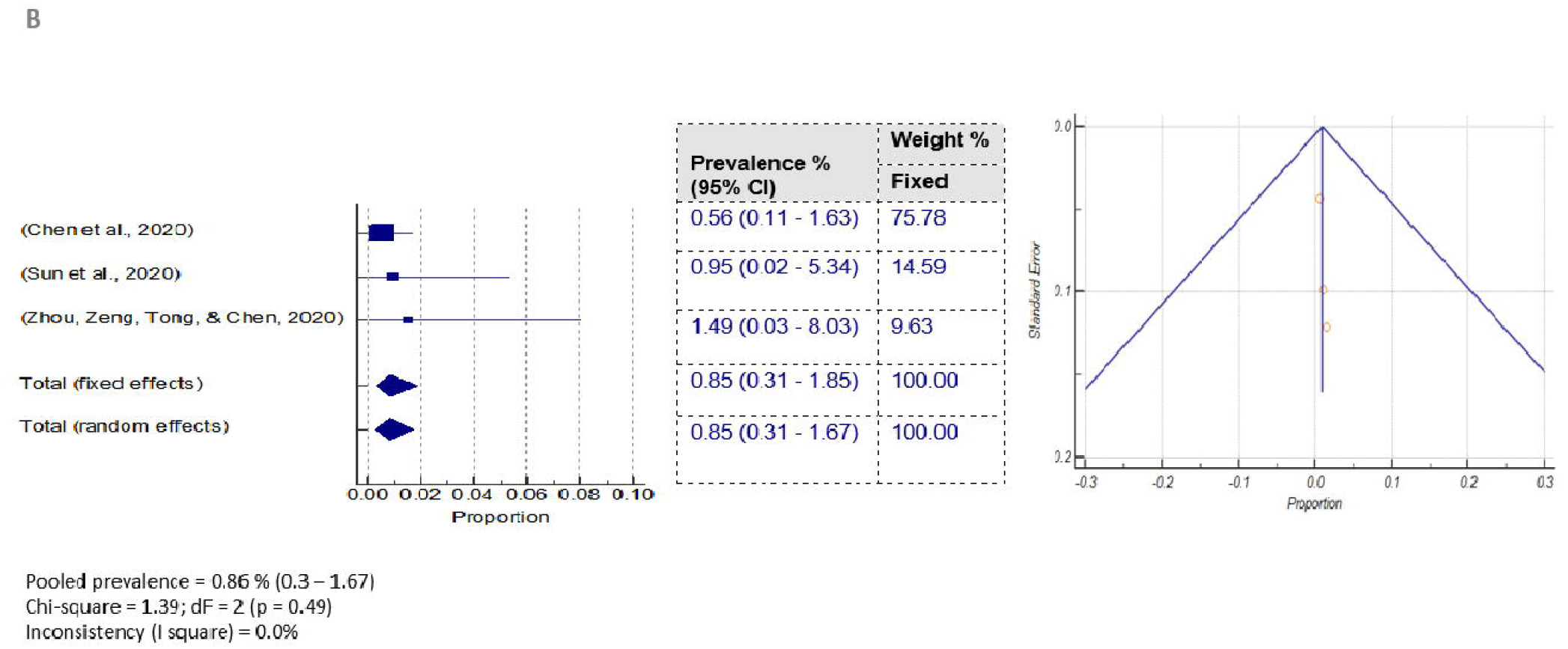
represents the Forest plot (left) and the Funnel plot (right) of the pooled prevalence of three studies which reported conjunctivitis as an initial symptom of the disease.

The study by *Wu et al*^10^ showed a much higher prevalence (26.31%) compared to the other studies. On removing this study from the forest plot, the I^2^reduced to 0.00% showing that this study contributed to much of the heterogeneity in this model.

### Conjunctivitis as an initial symptom of the disease

Conjunctivitis was reported as an initial symptom of the disease at the presentation by 3 studies (sample: 703 patients) in a total of 5 patients. As there was no significant heterogeneity between studies (I^2^= 0.0%, p=0.5), using the fixed-effects model the estimated pooled prevalence was 0.858 % (95% CI: 0.31-1.67). The forest plot with the estimated prevalence for each study has been presented in Figure 2B. All these 5 patients developed systemic symptoms and eventually diagnosed to have COVID-19 in subsequent days. There was no publication bias observed between studies (Figure 2B).

Apart from conjunctivitis, eye itching (in 1 patient, *Xu et al*^22^), epiphora (in 1 patient, *Wu et al*^10^), foreign body sensation (in 1 patient, *Karimi et al*^22^) and ocular secretions along with itching and dry eye (4 patients, *Hong et al*^19^)has been reported as an initial symptom of the disease, which didn’t progress further to any other ocular complications.

The time of presentation of conjunctivitis varied across studies and it has also been reported during the disease period^9,11,13,26^(in total 24 patients across studies and 1 case report^27^). The duration of systemic illness at the time of incidence of conjunctivitis ranged from 3-20 days. The conjunctivitis was usually self-healing^13,16^ and the resolution period ranged from 2 to 15 days^13,15,24-27^. However few studies reported the use of eye drops such as ganciclovir^13,15^, ribavirin^25^, valacyclovir^27^, antibiotics^13,25,28^ (ofloxacin, tobramycin, moxifloxacin) and artificial tears^13,24^ as supportive therapy for conjunctivitis.

*Chen et al*^13^ conducted a cross-sectional observational study in 534 diagnosed COVID-19 patients from two centers and found 25 patients (4.68%) with conjunctivitis. Additionally, they also found symptoms of dryness (20%), foreign body sensation (11.6%), tearing (50, 9.8%), itching (9.6%) and secretion (45, 8.8%), inpatients without conjunctivitis.

Similarly, in a questionnaire-based study conducted by *Hong et al*^*19*^ ocular symptoms was evaluated in 56 fully recovered discharged COVID-19 patients. The demographic details were obtained from hospital electronic medical records. The Salisbury Eye Evaluation Questionnaire (SEEQ) and Ocular Surface Disease Index (OSDI) were introduced by telephonic conversation. The patients were asked to recall any ocular symptoms experienced before, during, and after the onset of the disease. Fifteen patients (27%) reported ocular symptoms that include conjunctivitis, dry eyes, secretions, sore eye during the course of the disease. The SEEQ and OSDI scores were significantly higher post-illness.

Apart from the observational studies six case reports^23-28^ also described the incidence of conjunctivitis, of which, two reported conjunctivitis as an initial presentation of the disease, three reported conjunctivitis along with mild symptoms of cough, fever and sore throat as the first presentation followed by clinical signs and confirmed with laboratory investigations on subsequent days. The detailed characteristic has been shown in table 2. One report highlighted the occurrence of bilateral conjunctivitis in a 30-year-old gentleman on the 13^th^ day of illness. The typical presentation of conjunctivitis was accompanied by foreign body sensation, tearing, conjunctival follicles and an enlarged preauricular lymph node.^26^

**Table 2:**
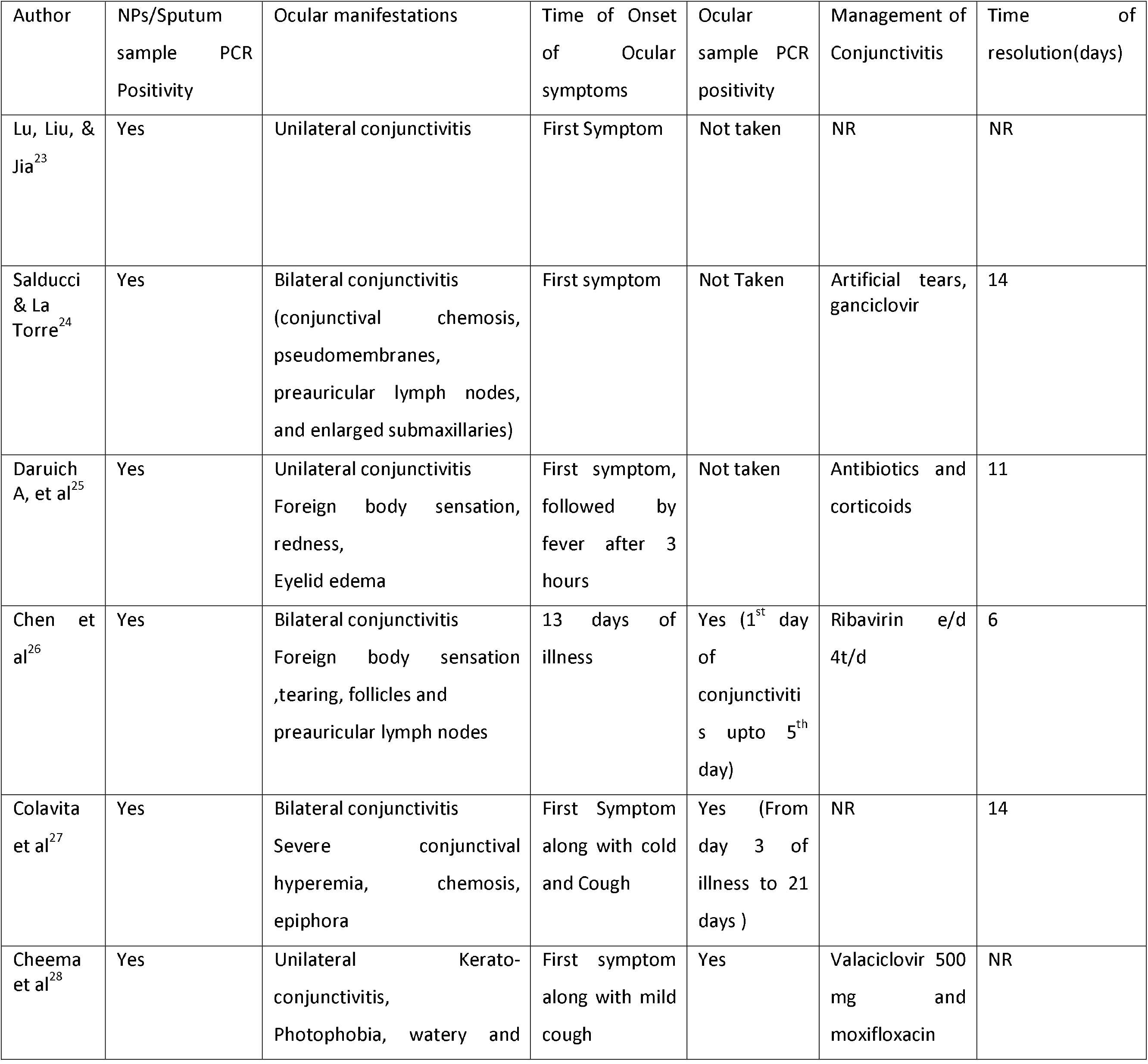
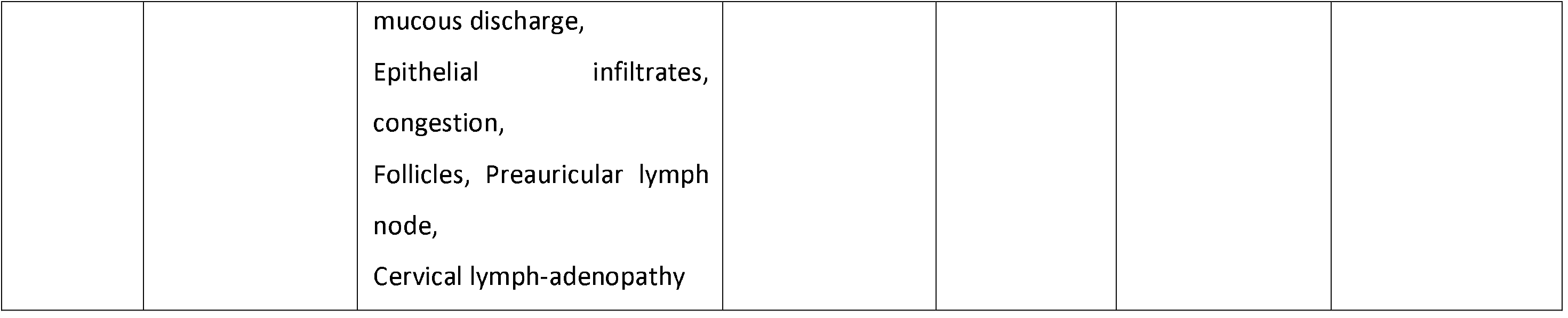
Characteristics of the included case-reports for the systematic review.

*Cheema et al*^28^ highlighted the first case presented as kerato-conjunctivitis characterized by multiple sub-epithelial infiltrates, corneal staining and follicular reaction of the conjunctiva. Initially, it was misdiagnosed and treated for epidemic kerato-conjunctivitis. Her ocular signs and symptoms worsened on subsequent follow-ups visists and on the 5^th^ day, NPs were positive for SARS-CoV-2 RNA.^28^

It is interesting to note that a 72-year-old gentleman developed bilateral conjunctivitis (conjunctival chemosis, pseudomembranes, preauricular lymph nodes and enlarged submaxillaries) as the only sign and symptom of COVID-19, while he was traveling on a cruise ship. He was tested positive for SARS-CoV-2 RNA in NPs. The patient had no systemic symptoms and approximately after 2 weeks his conjunctivitis resolved and he was tested negative twice in NPs .^24^

### The prevalence PCR positivity in NPs /Sputum samples and ocular samples

All the 14 studies have evaluated NPs or sputum samples for viral RNA detection by PCR method. However, 12 studies (sample: 667 patients) have collected NPs or sputum samples as well as ocular samples for PCR study. The estimated pooled positivity for NPs and sputum samples was 89.8% (95% CI: 78.80-79.0; random-effects model, I^2^= 93.63 %, p= <0.05) whereas for ocular samples was 2.90 % ((95% CI: 1.77 – 4.46; fixed-effect model, I^2^= 40.05%, p=0.07). Figure 3A and 3B represented the forest plot and the funnel plot with the estimated prevalence for each study for PCR positivity.

**Figure 3A.**
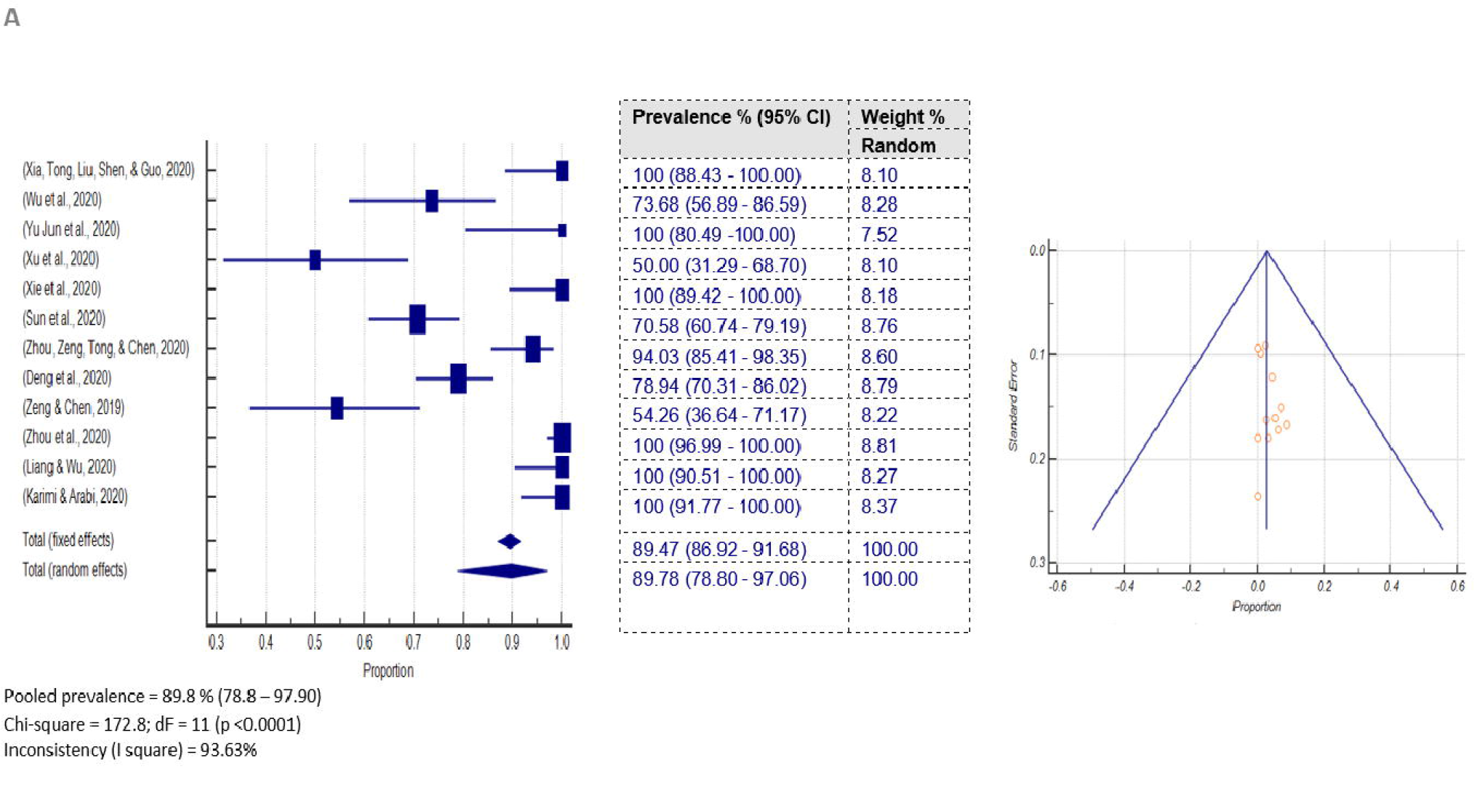
represents the Forest plot and Funnel plot of the pooled prevalence of PCR positivity in Nasopharyngeal/ Sputum samples.

**Figure 3B.**
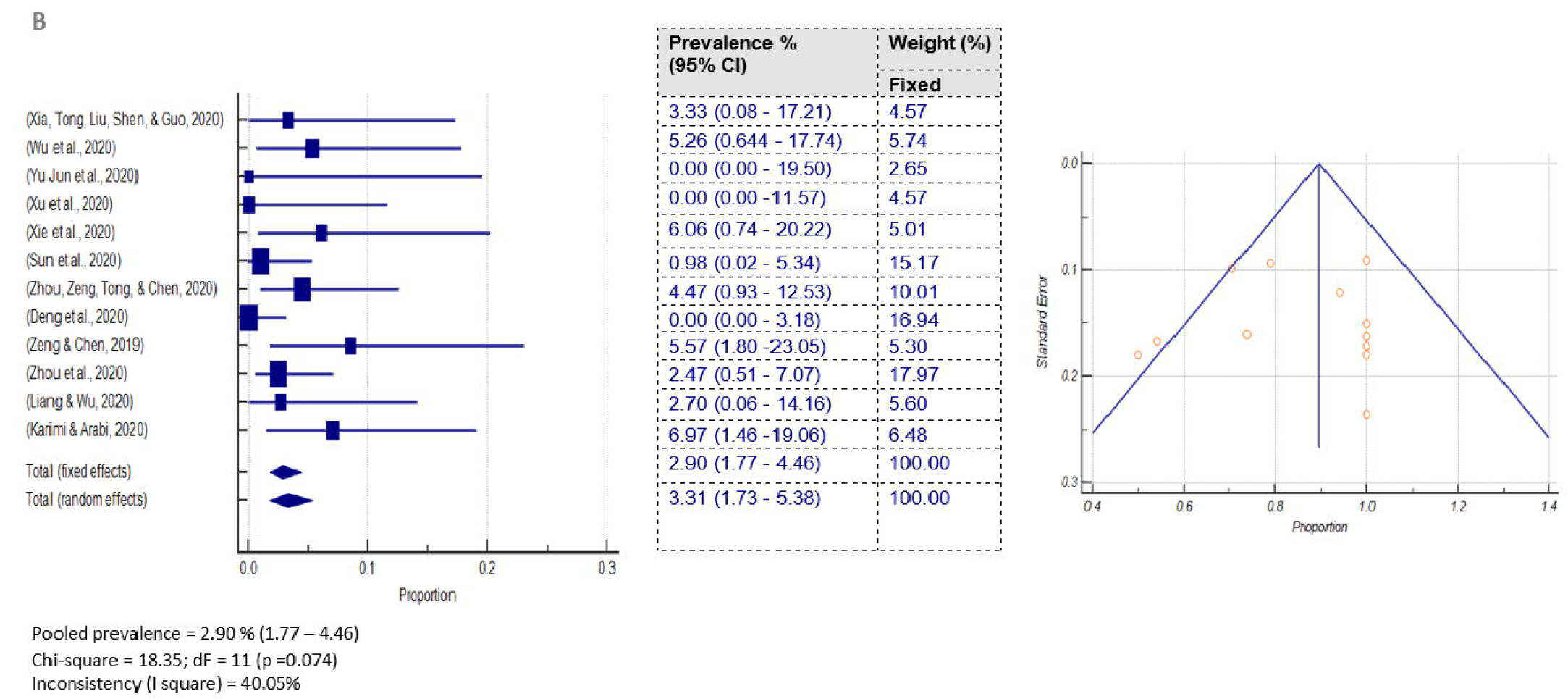
represents the Forest plot and Funnel plot of the pooled prevalence of PCR positivity in ocular samples.

#### Ocular samples positivity in conjunctivitis patients

A total number of 9 studies evaluated the presence of viral RNA in ocular samples in conjunctivitis patients. Based on the fixed-effects model (I^2^=13.15, p=0.32) results, the estimated pooled prevalence of viral RNA positivity in ocular samples of conjunctivitis patients was 32.74 % (95% CI: 17.49 – 51.23). The ocular sample positivity was also observed in asymptomatic patients. Figure 4 represented the Forest plot and Funnel plot of the pooled prevalence of viral RNA positivity.

**Figure 4.**
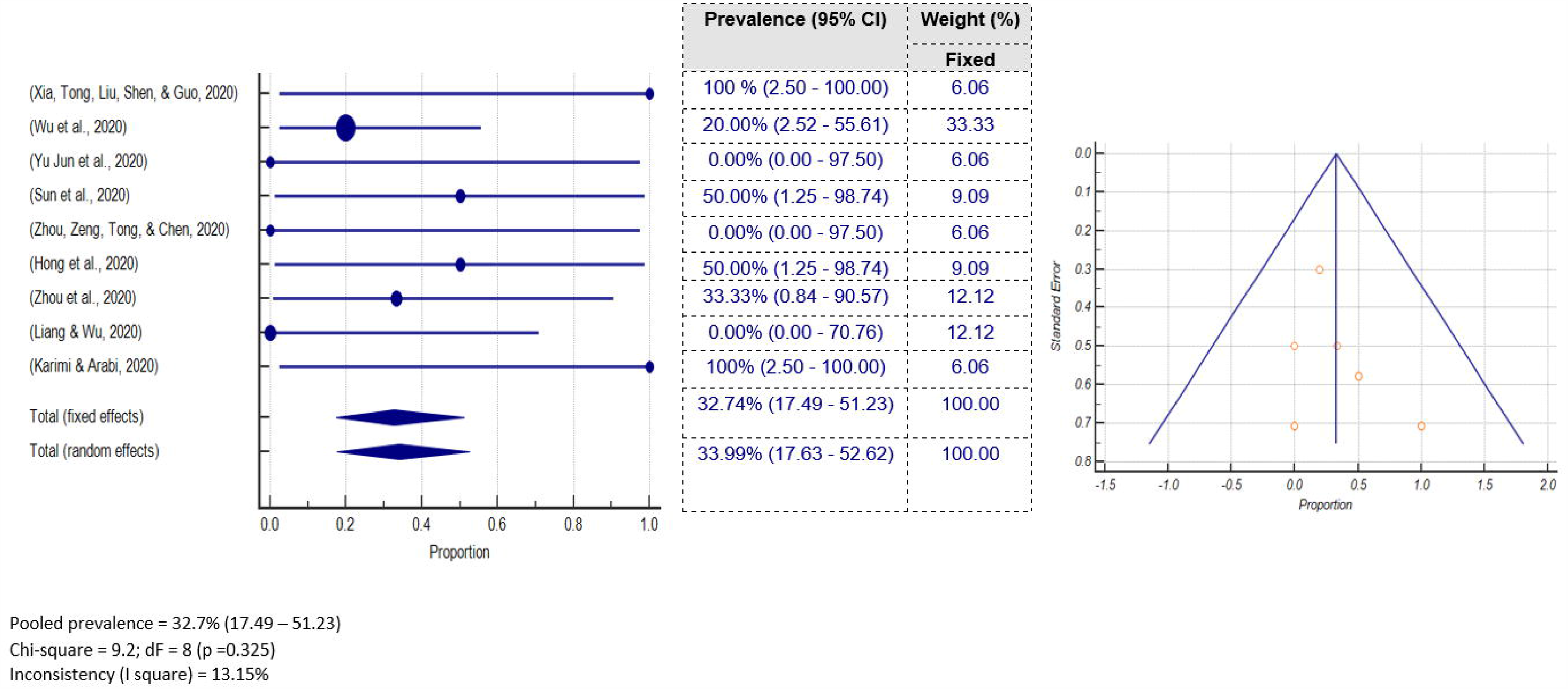
represents the Forest plot and Funnel plot of the pooled prevalence of viral RNA positivity in ocular samples of conjunctivitis patients.

In a total number of 24 patients with conjunctivitis, 7 patients (29 %) showed positive results in ocular samples by PCR assay. Whereas, 13 patients (54%) showed PCR positive only in their NPs/Sputum samples and 4 patients (16 %) were negative for both NPs/Sputum as well as ocular samples.

#### Time of collection of ocular samples

The time of ocular sample collection was reported by 9 studies. A detailed outline is presented in table 3. A maximum number of studies had collected NPs and ocular samples at the same time.

**Table 3:** Studies reported time for collection of Ocular sample PCR positivity results.

| Author | No. ocular PCR +ve<br>(sample size) | Time of collection<br>(days) | Time specifically for PCR<br>+ve patients (day) |
| --- | --- | --- | --- |
| *Xia et al*<sup>9</sup> | 1 (30) | 7.33±3.82 | 3rd |
| Yu Jun et al<sup>11</sup> | 0 (17) | 3-20<br>Repeated sample<br>collection | Nil |
| Xu et al<sup>12</sup> | 0 (30) | ≤ 7 | Nil |
| Xie et al<sup>14</sup> | 2 (33) | ≤7 | 5th and 4th |
| Sun et al<sup>15</sup> | 1 (102) | 18.15 ±7.57 | 3rd |
| Deng et al<sup>17</sup> | 0 (114) | 11 ± 6.3 | Nil |
| Yan et al<sup>18</sup> | 3 (35) | 9.71±6.50 | 8<sup>th</sup>, 7<sup>th</sup>, 29<sup>th</sup> |
| Zhou et al<sup>20</sup> | 3 (121) | 15±8.80 | NR |
| Karimi et al<sup>22</sup> | 3 (43) | 3.27 (range:1-7 days) | NR |

*Yu Jun et al*^*11*^ in a prospective study evaluated tear samples collected from 17 confirmed COVID-19 patients every week from day 3 of symptoms to day 30. They were analyzed using PCR assay, which yielded no positive results in all the cases at all weeks. However, one patient developed conjunctival congestion and chemosis during the hospital stay but still showed no positivity in the tears.

Similarly, in another prospective interventional case series, where ocular samples were collected twice in an interval of 2 to 3 days in 30 confirmed COVID-19 cases yielded RT-PCR positivity in only 1 patient with conjunctivitis.^12^

## Discussion

In this study, we performed a comprehensive systematic review and meta-analysis to provide an in-depth outline of ocular manifestations among patients with COVID-19.

Based on our meta-analysis, the prevalence of conjunctivitis/ conjunctival congestion was 5.17 % (95% CI: 2.90-8.04). Additionally, we also found that conjunctivitis, as a presenting manifestation of COVID-19, is seen in <1% cases (0.858 % ((95% CI: 0.31-1.67)).The reported clinical characteristics of conjunctivitis mostly resemble the features of viral conjunctivitis. The associated features include itching, epiphora, secretion, chemosis, foreign body sensation, follicular reactions of the palpebral conjunctiva and few reported extra-ocular manifestations such as lymphadenopathy. Although some patients without conjunctivitis also had mild self-limiting ocular symptoms like epiphora, itching and foreign body sensations. As reported by 2 studies dryness is also one of the associated features.^13,19^ The first ocular symptom was reported by Wang Guangfa (national expert on pneumonia) who experienced conjunctivitis as the first symptom of the disease after he returned from Wuhan.^23^ He believed that he had developed conjunctivitis because of not using any protective eyewear during the inspection. He subsequently developed a fever and tested positive for SRAS-CoV-2 RNA in NPs After this incident, there were an increased awareness and interest to investigate the ocular manifestations and transmission of the virus in the scientific world. In the later days, there were case reports and observational studies published reporting conjunctivitis as an initial presentation of the disease and also during the course of the disease. The incidence of ocular symptoms was observed in various stages of the diseases ranged from the early latent stage (day 0) without any systemic symptoms or even a negative NPs to the late stage of the disease (day 20).

In contrast to the published literature, the presentation of conjunctivitis has been reported in patients affected with the less pathogenic human coronavirus NL63 (HCoV-NL63).^28,29^Conjunctivitis was first observed as a presenting symptom of the disease in a 7-year-old child along with fever and bronchiolitis.^30^ A retrospective study conducted by Vabret et al ^29^ in France analyzed 18 children’s with upper respiratory tract illness due to HCoV-NL63 found conjunctivitis in 3 (17%) children. So far, there are no reports on ocular complications published for SARS-CoV-1, MERS-CoV.

From the observations, it was clear that ocular manifestations were not so common in patients with COVID-19. Conjunctivitis is the only ocular manifestation of the disease observed so far.

The efficacy of the use of antiviral agents like ganciclovir, ribavirin, valaciclovir in treating conjunctivitis caused by SARS-CoV’s had not been evaluated. However, the use of antibiotics and artificial tears can be used to reduce the chances of secondary bacterial infections and symptomatic relief respectively.

In our study, the meta-analysis results of ocular samples positivity among COVID-19 was low around 2.90 % (95% CI: 1.77 – 4.46), whereas the positivity rate for NPs and sputum was 89.8% (95% CI: 78.80-79.0). Most of the studies have collected NPs and ocular simultaneously at the same time. Our observations demonstrate that ocular samples can be positive at any time point during the illness period. In a case report by *Colavita et al*^*27*^, an ocular sample from a conjunctivitis patient yielded positive results from day 3^rd^ to day 21 of illness. While the conjunctivitis was apparently healed, the patient again showed PCR positivity in ocular sample on the 27^th^ day even after it became undetectable in the NPs. Similar observations were noted by *Yan et al*^18^ in a patient without conjunctivitis on the 29^th^ day of illness. Patients without conjunctivitis also showed viral RNA in their ocular samples and about 16% of conjunctivitis patients were negative in both NPs/sputum as well as ocular samples.

Looking back, after the breakout of SARS-CoV-1 in 2003, a case series published had reported the detection of SARS-CoV RNA in the conjunctival samples in 3 (37%) out of 8 confirmed SARS patients in their early phase of the disease. One female healthcare worker yielded SARS positivity only in the tears.^31^ Whereas, in a prospective interventional case series none out of 17 patients showed viral RNA positivity in their conjunctival scrapings.^32^ The discrepancy might be due to factors like low viral load in conjunctival secretion, time of collection of ocular, the sensitivity of the PCR kits, method of collection of, and also the influence or interaction of antiviral systemic medications administered.

The ocular surface poses receptors for several respiratory viruses. The receptors of some species of adenovirus and avian influenza (α-2-3-linked sialic acid,CD46, desmoglein-2) and the human influenza virus (α-2-6-linked sialic acid) are abundantly expressed in the corneal, conjunctival epithelium as well as nasal and tracheal mucosal lining.^5,6,33^ The similarities in the distribution of cellular receptors in the ocular surface mucosa and the respiratory tract pose a high risk of viral tissue tropism adherence and transmission.^33^ The binding cellular receptor, angiotensin-converting enzyme 2 (ACE2) for SARS-CoV-1 and SARS-CoV-2 and the HCoV-NL63 is extensively expressed in the lung alveoli, heart, testes and intestinal epithelial cells. In the eye, ACE2 expressions are found on the corneal, conjunctival epithelium, aqueous and vitreous humor although in fewer concentrations as compared to lungs and gastrointestinal tissues.^5,6^

The current evidence suggests the binding of SARS-CoV Spike protein (S240) with the ACE2 receptor is amplified by heparin sulfate and TMPRSS2 proteins within the host cell.^34^ So far, the presence of TMPRSS2 proteins has not been reported on the ocular surfaces. Furthermore, the anti-microbial activity of the tears can play a protective role in the host defense mechanisms by the process of viral elimination. The presence of lactoferrin, lipocalin 2, and immunoglobin A in the tears can inhibit viral attachment by interfering with ACE2 receptors. There is a 150 fold increment in the levels of lactoferrin in SARS CoV patients.^35,36^

Although the fact that the proteins crucial for the renin-angiotensin-aldosterone system (RAAS) expressed in many ocular structures like aqueous, pigmented epithelium, and the retina has been proved^29^.The expression of such proteins both in the conjunctival and corneal epithelium is less, which could also be a reason for the discrepancy in the number of individuals to show PCR negativity in ocular despite a positive NPs by PCR.

In experimental animal models, such as the feline coronavirus (feline infectious peritonitis) and the murine coronavirus (mouse hepatitis virus), apart from viral conjunctivitis are also capable of producing sight-threatening conditions like granulomatous anterior uveitis, retinal vasculitis, choroiditis, retinal detachment, virus-induced macular degeneration and optic neuritis in cats with poor prognosis.^1^ Moreover, about 90% of conjunctival isolates were tested positive.

In a recent study, rhesus macaques, when inoculated with infectious doses of SARS-CoV-2 through the conjunctival sac became potentially infected with the virus. Viral load was detectable in the throat, nasal and conjunctival samples post day 1 of inoculation and its distribution was detected in various anatomical locations such as lacrimal gland, nasolacrimal system, optic nerve, alimentary tract, lower lobe of the lungs. Radiologic investigations showed interstitial pneumonia. The authors concluded according to the observations of the study the possibility of infection of SARS-CoV-2 via the conjunctival route.^7,37^

The possible hypothesis for the presence of the virus on the ocular surface and transmission of the virus through the ocular route can be drawn in several ways: Firstly, an exposed ocular surface can directly inoculate the virus from the external environment through contaminated droplets, aerosols or an infected hand to eye contact. The transfer of microorganisms along with the tears from the ocular surface into the nasal cavity and the upper part of respiratory tissues is because of the continuation of the mucous membrane via the puncta into the nasolacrimal duct and the nasopharyngeal space. This can eventually deliver the virus into the upper respiratory tissues and the gastrointestinal tract when swallowed. Secondly, absorption and exudation from the ocular tissues like conjunctiva and corneal epithelial, lining of the lacrimal system epithelial cells and the inner lining of the pharynx are also possible. Thirdly, the virus can also travel retrograde into the nasal and ocular fluids.

We came across few limitations in the studies: 1) The Majority of the studies had a small sample size, except for a few, 2) The time of ocular sampling differed among each study 3) The substantial heterogeneity can influence the results in few cases. 4) The study was carried-out during the peak stage of the on-going pandemic; therefore less serious conditions including ocular complications may be overlooked.

## Conclusion

This review provides a comprehensive characterization of clinical features and the ocular symptoms experienced by patients with COVID-19. Conjunctivitis may present as the first symptom of the disease making the patient seek medical care at the earliest. Our analysis showed the prevalence of conjunctivitis and ocular PCR positivity among COVID-19 patients was low indicates that the eye is a less affected organ. Although the ACE2 receptors are present on the ocular surface, the binding efficiency of the virus might be affected by the absence of some other proteins and anti-microbial activity of the tears. Ocular symptoms can manifest at any point in the disease course. However, viral shedding in the tears has been rarely isolated compared to nasopharyngeal or Sputum samples. Due to the inconsistent evidence of the virus in the ocular samples its ability to infect the ocular structures, spread through ocular fluids, or use of ocular as a diagnostic tool is not clearly understood.

## Data Availability

This is a systematic review and Meta-analysis.

